# Association of optic disc parameters and glaucoma incidence based on automated segmentation, evidence from the UK Biobank

**DOI:** 10.1101/2023.11.06.23298106

**Authors:** Pinxuan Zhu, Shuang He, Danli Shi, Mingguang He

## Abstract

**Objective:** To assess the correlation between glaucoma incidence and optic disc parameters obtained through an automated deep learning (DL) algorithm segmentation.

**Methods and Analysis:** We obtained eligible fundus photographs and corresponding participant data from the UK Biobank. To accurately assess the optic disc parameters and their relationship with glaucoma incidence using Cox proportional hazard regression models, we developed a DL algorithm that automatically segmented the optic disc and cup and calculated various parameters including the vertical cup-to-disc ratio (VCDR), ovality index, cup-to-disc area ratio, rim area, disc area, and disc rotation from the fundus photos. We performed two logistic regression models, with model A comprising sociodemographic and health covariates and model B including additional ophthalmic features. Receiver operating characteristic curves (ROC) and areas under the curve (AUC) were plotted and calculated for each model to evaluate their performance.

**Results:** A total of 44,376 subjects with fundus photos were included in our study. After a median follow-up of 10.1 years, 354 incident glaucoma were documented. Subjects with larger VCDR had a higher risk of incident glaucoma; the HR (95% CI) was 2.05 (1.57-2.66) in the multivariable-adjusted model (p<0.001). The results remain significant in the sensitivity analysis that excluded fundus photographs with “Reject” quality. After adding the optic disc parameters into the regression model A, the AUC increased by 4.2% to 78.6%.

**Conclusion:** The VCDR calculated by automatic optic disc segmentation model shows potential as a biomarker for evaluating the risk of glaucoma.

**What is already known on this topic:** Glaucoma is a worldwide leading cause of irreversible vision loss, and its early diagnosis is of great necessity.

**What this study adds:** Data from the UK Biobank shows the optic disc parameters and their relationship with glaucoma incidence.

We develop a DL-based algorithm for optic disc segmentation in Color fundus photos and validate its efficacy in glaucoma prediction.

**How this study might affect research, practice or policy:** The VCDR calculated using an automatic optic disc segmentation based on a DL model can serve as a biomarker to predict the incidence of glaucoma.

## Introduction

Glaucoma is a leading cause of vision impairment, affecting over 60 million people worldwide. This number is projected to increase to 111.8 million by 2040, with a disproportionate impact on populations in Asia and Africa.^1,2^ Symptoms of the condition include the loss of retinal ganglion cells, degeneration of the optic nerve, and defection of the visual field.^3^ Due to the absence of visible symptoms in the early stages, up to half of the patients may remain undiagnosed. Therefore, it is crucial to detect the disease at an early stage to prevent further deterioration.^4,5^

Structural alterations in the optic disc are the primary factor in diagnosing glaucoma.^6-8^ The vertical cup-to-disc ratio (VCDR), cup-to-disc area ratio, and the ISNT (inferior ^≥^superior ^≥^nasal ^≥^ temporal) rule for rim width pattern of the optic disc are frequently used to aid in the diagnosis of glaucoma.^9-12^ However, it is crucial to acknowledge that not all modifications in the optic disc are indicative of glaucoma, as some alterations may be due to physiological factors or other ocular conditions.^13-17^ Furthermore, diagnosing glaucomatous optic neuropathy (GON) is reliant on subjective and empirical assessments, which adds to the complexity of diagnosing glaucoma.^18^ Color fundus photos (CFPs) are a valuable tool in ophthalmology as they allow for the visualization of lesion characteristics in a non-invasive, accessible, and cost-effective manner, making it a widely used method for large-scale detection.^19,20^ The advent of deep learning (DL) algorithms has revolutionized the field of ophthalmology by providing automatic methods to segment and analyze optic disc parameters from fundus images, thus mitigating the bias associated with subjectivity^21-23^. However, previous studies investigating the association between glaucoma incidence and optic disc parameters have largely relied on manual labelling methods to obtain disc parameters, resulting in limited sample sizes and specific glaucoma populations^6,8^.

Therefore, we aim to develop a DL-based algorithm for optic disc segmentation in CFPs and validate its efficacy in glaucoma prediction.

## Materials and Methods

### Study population

UK Biobank is a large-scale biomedical database containing more than 500,000 participants aged 40-69 across England, Scotland, and Wales during 2006-2010.^24^ Over 60,000 people took fundus photographs at baseline using Topcon 3D OCT 1000 Mk2 instrument. Baseline information on sociodemographic, lifestyle and health-related factors was collected through questionnaires at recruitment, and health-related outcomes were obtained with participant consent through regular contact with UK health and medical records such as inpatient data and death registers. Physical measures including eye measurements were obtained at the baseline as well. Patients with glaucoma at baseline (judging by the time of glaucoma and those with IOP higher than 21mmHg or VCDR larger than 0.8) were excluded. A flow diagram for participant inclusion is illustrated in Figure S1. In brief, after excluding patients without fundus images and those with baseline glaucoma, as well as patients with missing data on systemic or ocular variables, a total of 44,376 participants with fundus images were enrolled in the study.

All participants provided written informed consent, and the study was approved by the UK Biobank’s research ethics committee, endorsed by the National Research Ethics Service (reference 11/NW/0382) and the Research Tissue Bank approval.

### Study database

In this study, we utilized six publicly available glaucoma databases that offer pixel-level segmentation of the optic disc and optic cup. These databases include 1200 images from the REFUGE challenge dataset, 1020 images from the G1020 dataset, 693 images from the RIGA dataset, 159 images from the RIM-ONE dataset, 101 images from the DRISHTI-GS dataset, and 100 images from the GAMMA dataset.

The REFUGE challenge dataset is composed of fundus images from several hospitals and clinics in two different sizes (2124 × 2056 pixels and 1634 × 1634 pixels).^25^ Optic disc and cup annotations are performed on each image in the original database. The G1020 contains images of 432 patients collected in Germany with a 45-degree field of view after the use of dilated drops.

The images’ size is between 1944 × 2108 and 2426 ×3007 pixels.^26^ The RIGA consists of photos taken from Messidor dataset and two clinics in Saudi Arabia which provide boundary annotations of optic disc and cup.^27^ The RIM-ONE consists of high-resolution fundus photos collected in Spanish with 21.1 megapixels. The segmentation of the optical disc and cup for each photo is provided.^28^ The DRISHTI-GS dataset is composed of images centered on optic disc with a field-of-view of 30-degrees and of dimension 2896 × 1944 pixels. The GAMMA dataset contained both 2D fundus color images and 3D Optical Coherence Tomography (OCT) volume. The fundus photos in GAMMA dataset provide the mask of the cup and optic disc with a resolution of 2000 × 2992 pixels or 1934 × 1956 pixels.^29^

We split the images of each dataset into training, validation and test sets in an 8:1:1 ratio. In total, 3273 images are used for the experiment.

### Deep learning algorithm

We used the pix2pixHD algorithm, a generative adversarial network (GAN) capable of generating high-resolution images and performing image semantic editing, to segment the optic disc and optic cup in the fundus photos.^30^ The generator in this model learns the input in a coarse-to-fine manner, and the discriminator assesses the authenticity of the generated images across multiple scales. The images were first cropped to ensure that only the field of view was included before being input into the model. During training, the batch size was set to 1, and the learning rate was 0.0002. A total of 100 training epochs were run, and the model with the highest Dice index on the validation set was selected. To evaluate the segmentation performance, we computed accuracy, sensitivity, specificity, and F1 score for each dataset.

### Disc-cup measurement

We applied established formulas to the segmented images to obtain various optic disc parameters. The vertical cup-to-disc ratio (VCDR) was calculated as the ratio of the vertical diameter of the cup to the vertical diameter of the disc on the vertical line. The ovality index was the quotient of the smallest and largest optic disc diameters. We quantified the disc and rim areas in pixels. The cup-to-disc area ratio was defined as the ratio of the cup area to the disc area. We used the optic disc rotation degree, the angle between the longest diameter and the vertical line, to determine the disc rotation. Negative disc rotation values for the right eye denoted superior rotation.

### Acertainment of glaucoma

We identified glaucoma cases based on UK Biobank inpatient hospital data from 1998, using International Classification of Diseases tenth Revision (ICD10) codes.^31^ We also included participants who reported glaucoma in the follow-up questionnaires, used glaucoma medication, or had a history of glaucoma surgery between 2010 and March 31, 2021.

### Covariates

We included covariates including, sociodemographic information, involving age, sex(female/male), ethnicity(non-whites/whites), body mass index (BMI), smoking status(never/former smoker/current smoker), systolic and diastolic blood pressure, intraocular pressures, spherical equivalent for both eyes, metabolic biomarkers comprising cholesterol, high-density lipoprotein (HDL), and low-density lipoprotein (LDL), history of hypertension(no/yes), history of diabetes mellitus (DM) (no/yes), history of myopia (no/yes).

We collected data on these covariates from baseline questionnaires and measurements. We used UK Biobank inpatient data on hospital admissions before recruitment to determine the history of hypertension and DM. History of myopia was identified via self-reported data from UK Biobank. To minimize the confounding impact, we introduced these factors as potential confounders into models.

### Statistical analysis

We reported the baseline characteristics of the study participants using descriptive statistics, such as means, standard deviations (SDs), numbers, and percentages. ANOVA and t test were used to compare the baseline data between glaucoma and normal population. We used Cox proportional risk models to assess the association between optic disc parameters and incident glaucoma. We adjusted the models for the following covariates: baseline age, sex, and ethnicity (model I); BMI, smoking status (never, ever, or unknown), history of hypertension (yes or no), history of diabetes (yes or no), HDL, LDL, Cholesterol, DBP, SBP (model II); IOP, SE and HM status (model III). To verify the validity of our results, we performed a sensitivity analysis using high quality images.

In addition, we developed two logistic models using age, sex, ethnicity, BMI, HDL, LDL, cholesterol, DBP, SBP, hypertension, diabetes, smoking, IOP, SE and HM as model A, additional optic disc parameters (VCDR, ovality index, cup to disc area ratio, rim area, disc area and disc rotation) as model B. Receiver operating characteristic curves (ROC) and areas under curves (AUC) of each model were plotted and calculated.

A set of sensitivity analyses was performed. Considering that the quality of fundus photography can affect the measurement accuracy of optic disc parameters, we analyzed whether this association would change if only individuals with “usable” and “good” photo quality were selected.

All statistical tests described here were two-sided, and differences at P < 0.05 were accepted as statistically significant. Statistical analyses were conducted using Stata V.15.0 software (StataCorp, College Station, Texas, USA) and R software (version 4.0).

## Results

After excluding 437,310 subjects without fundus photos, 8,368 subjects with baseline glaucoma and 12,453 subjects with missing systemic or ocular parameters, a total of 44,376 subjects were included in our study. A flow diagram outlining the participant inclusion process can be found in Figure S1.

In Figure 1A, we present an example of the segmentation results, while Figure 1B demonstrates the disc-cup measurements. Table S1 displays the segmentation performance of our deep learning model on various databases. Our segmentation algorithm demonstrated high accuracy, with a value of 0.999 across all datasets. The sensitivity ranged from 0.830 to 0.977, and specificity reached 0.999 to 1.00. The optic cup segmentation performance was generally better than that of the optic disc, as indicated by F1 scores ranging from 0.808 to 0.978.

**Figure 1.**
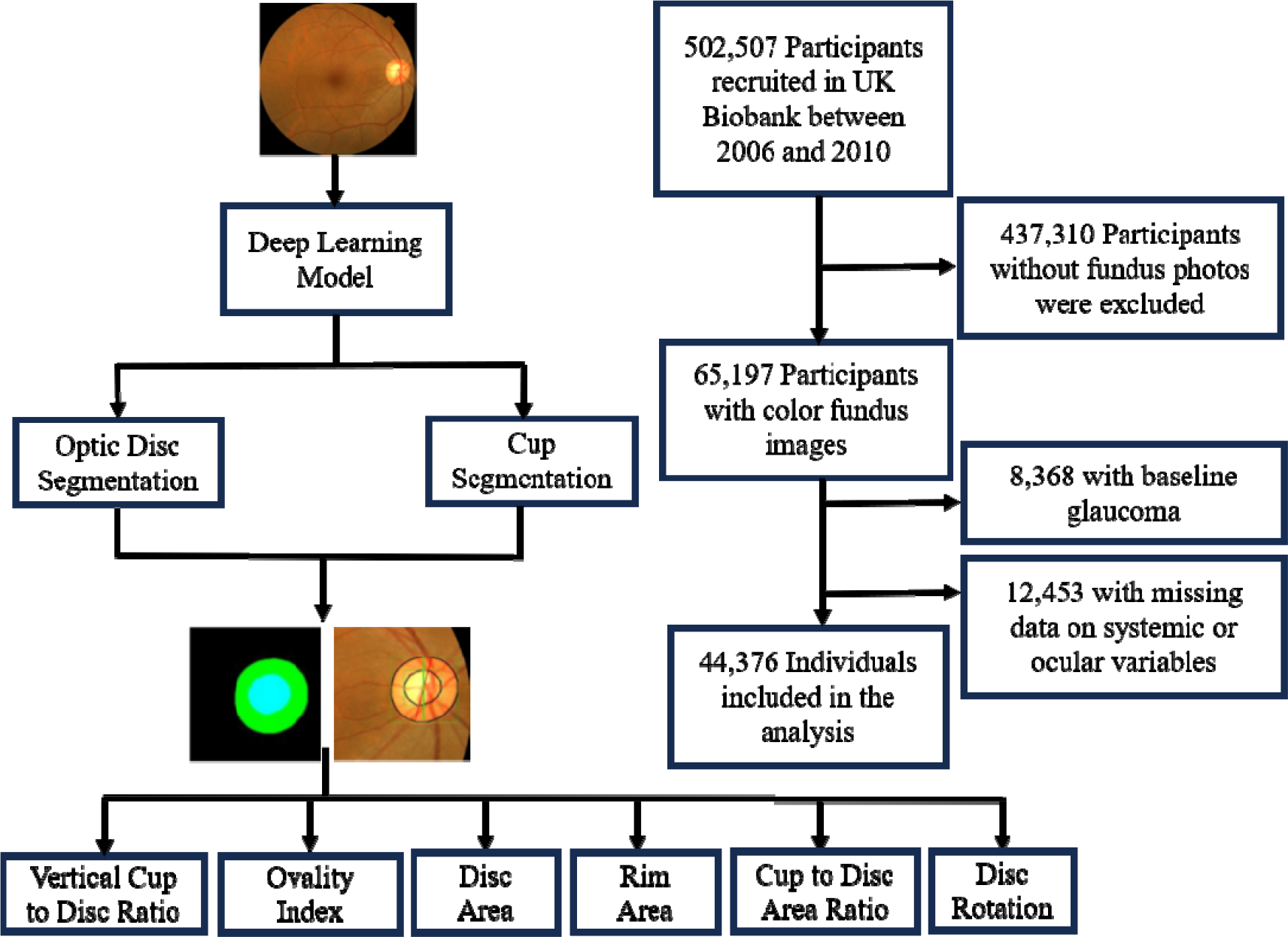
Flow chart of the study.

Table 1 presents the baseline characteristics of the 44,376 participants included in the study. The mean age of the participants was 56.21 ± 8.21 years, with 44.91% being males. There were significant differences in age, sex, BMI, ethnicity, presence of hypertension and diabetes, SE, and presence of high myopia (HM) between participants who developed incident glaucoma and those who did not (P < 0.05 for all).

**Table 1.** Baseline characteristics of participants in the UK Biobank.

| Characteristics | Mean (SD)/ n (%) |  |  | P value |
| --- | --- | --- | --- | --- |
|  | No glaucoma<br>(N=44,022) | Glaucoma<br>(N=354) | All<br>(N=44,376) |  |
| Age | 56.17(8.21) | 61.06(6.17) | 56.21(8.21) | <0.001<sup>*</sup> |
| Gender(Male,%) | 19,747(44.86%) | 184(51.98%) | 19,931(44.91%) | 0.007<sup>†</sup> |
| BMI, kg/m<sup>2</sup> | 27.26(4.70) | 28.00(5.12) | 27.26(4.70) | 0.018<sup>*</sup> |
| Ethnic(Whites,%) | 39,738(90.27%) | 308(87.01%) | 40,046(90.24%) | 0.039<sup>†</sup> |
| HDL, mmol/L | 1.49(0.39) | 1.43(0.39) | 1.49(0.39) | 0.883<sup>*</sup> |
| LDL, mmol/L | 3.54(0.85) | 3.44(0.86) | 3.54(0.85) | 0.935<sup>*</sup> |
| Cholesterol,<br>mmol/L | 5.69(1.12) | 5.54(1.15) | 5.69(1.12) | 0.568<sup>*</sup> |
| Systolic Blood<br>Pressure, mm Hg | 136.00(18.10) | 137.38(17.64) | 136.01(18.09) | 0.506<sup>*</sup> |
| Diastolic Blood<br>Pressure, mm Hg | 81.50(9.97) | 80.86(9.76) | 81.50(9.97) | 0.577<sup>*</sup> |
| Hypertension | 22,163(50.35%) | 197(55.65%) | 22,360(50.39%) | 0.047<sup>†</sup> |
| Diabetes | 2,421(5.50%) | 34(9.60%) | 2,455(5.53%) | 0.001<sup>†</sup> |
| Smoking | 4,088(9.29%) | 29(8.19%) | 4,117(9.28%) | 0.480<sup>†</sup> |
| IOP, mmHg | 14.96(2.91) | 15.70(3.03) | 14.97(2.91) | 0.302<sup>*</sup> |
| SE, D | -0.37(2.64) | -0.79(3.52) | -0.37(2.64) | <0.001<sup>*</sup> |
| HM | 1,683(3.82%) | 28(7.91%) | 1,711(3.86%) | <0.001<sup>†</sup> |

SD: Standard Deviation; BMI: Body Mass Index; HDL: High Density Lipoprotein; LDL: Low Density Lipoprotein; IOP: Intraocular Pressure; SE: Spherical Equivalent; HM: High Myopia.
<sup>\*</sup> ANOVA test. <sup>†</sup> Chi-squared test.

We used Cox proportional hazard models to assess how optic disc segmentation parameters relate to incident glaucoma (Table 2). We found that larger VCDR increased the risk of incident glaucoma significantly (multivariable-adjusted HR = 2.05, 95% CI: 1.57-2.66, p<0.001). None of the other parameters had a significant effect. When excluding 12,686 fundus photographs with “Reject” quality (Table S2), the results kept robust for larger VCDR still predicted incident glaucoma significantly (multivariable-adjusted HR = 2.98, 95% CI: 2.07-4.31, p<0.001).

**Table 2.**
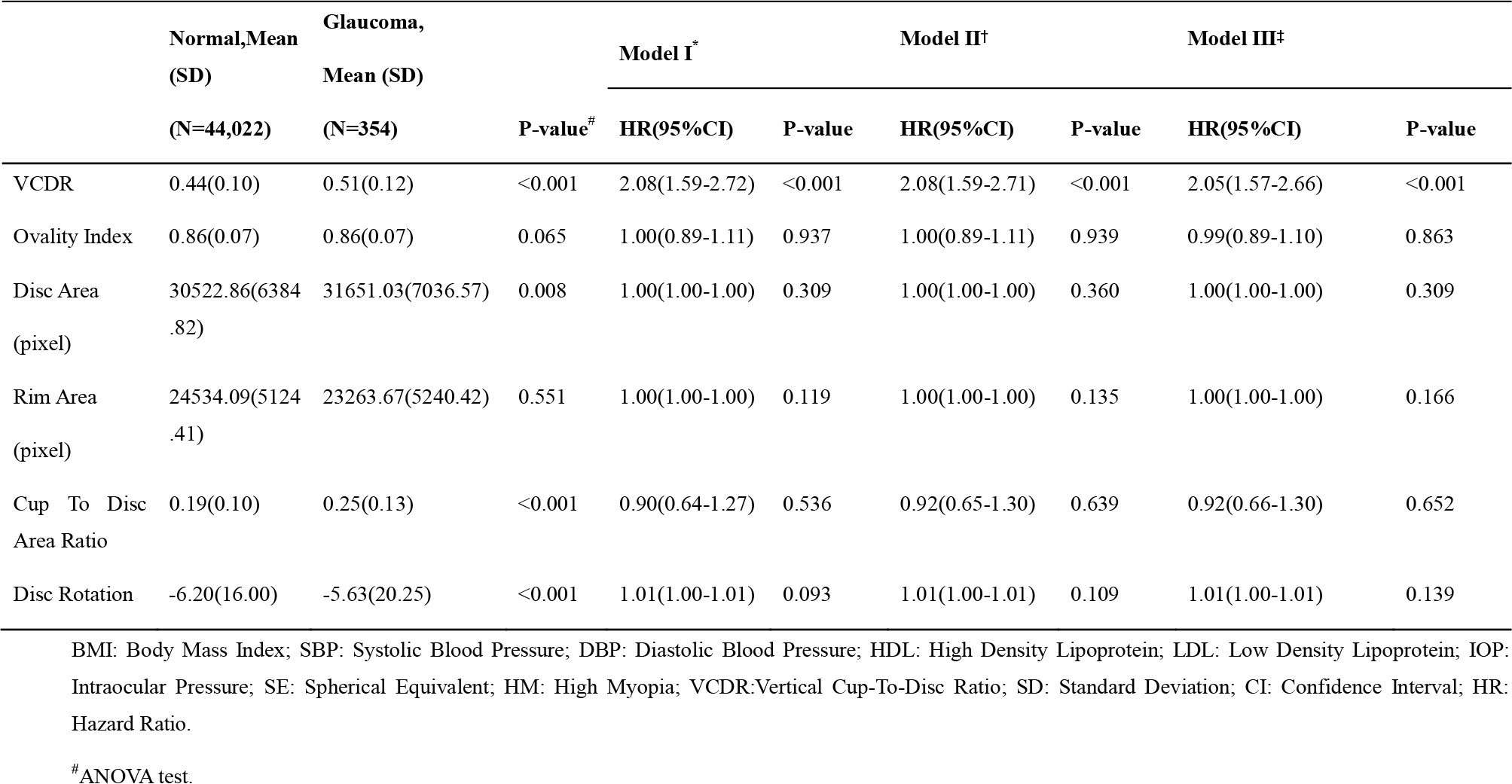
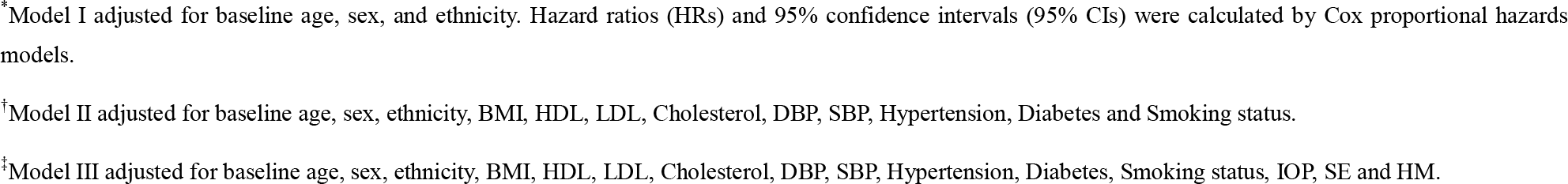
The association of optic disc parameters and incident glaucoma.

|  | Normal,Mean<br>(SD) | Glaucoma,<br>Mean (SD) |  | Model I<sup>*</sup> |  | Model II<sup>†</sup> |  | Model III<sup>‡</sup> |  |
| --- | --- | --- | --- | --- | --- | --- | --- | --- | --- |
|  | (N=44,022) | (N=354) | P-value<sup>#</sup> | HR(95%CI) | P-value | HR(95%CI) | P-value | HR(95%CI) | P-value |
| VCDR | 0.44(0.10) | 0.51(0.12) | <0.001 | 2.08(1.59-2.72) | <0.001 | 2.08(1.59-2.71) | <0.001 | 2.05(1.57-2.66) | <0.001 |
| Ovality Index | 0.86(0.07) | 0.86(0.07) | 0.065 | 1.00(0.89-1.11) | 0.937 | 1.00(0.89-1.11) | 0.939 | 0.99(0.89-1.10) | 0.863 |
| Disc Area<br>(pixel) | 30522.86(6384.82) | 31651.03(7036.57) | 0.008 | 1.00(1.00-1.00) | 0.309 | 1.00(1.00-1.00) | 0.360 | 1.00(1.00-1.00) | 0.309 |
| Rim Area<br>(pixel) | 24534.09(5124.41) | 23263.67(5240.42) | 0.551 | 1.00(1.00-1.00) | 0.119 | 1.00(1.00-1.00) | 0.135 | 1.00(1.00-1.00) | 0.166 |
| Cup To Disc<br>Area Ratio | 0.19(0.10) | 0.25(0.13) | <0.001 | 0.90(0.64-1.27) | 0.536 | 0.92(0.65-1.30) | 0.639 | 0.92(0.66-1.30) | 0.652 |
| Disc Rotation | -6.20(16.00) | -5.63(20.25) | <0.001 | 1.01(1.00-1.01) | 0.093 | 1.01(1.00-1.01) | 0.109 | 1.01(1.00-1.01) | 0.139 |

BMI: Body Mass Index; SBP: Systolic Blood Pressure; DBP: Diastolic Blood Pressure; HDL: High Density Lipoprotein; LDL: Low Density Lipoprotein; IOP: Intraocular Pressure; SE: Spherical Equivalent; HM: High Myopia; VCDR:Vertical Cup-To-Disc Ratio; SD: Standard Deviation; CI: Confidence Interval; HR: Hazard Ratio.
<sup>#</sup>ANOVA test.
\* Model I adjusted for baseline age, sex, and ethnicity. Hazard ratios (HRs) and 95% confidence intervals (95% CIs) were calculated by Cox proportional hazards models.
† Model II adjusted for baseline age, sex, ethnicity, BMI, HDL, LDL, Cholesterol, DBP, SBP, Hypertension, Diabetes and Smoking status.
‡ Model III adjusted for baseline age, sex, ethnicity, BMI, HDL, LDL, Cholesterol, DBP, SBP, Hypertension, Diabetes, Smoking status, IOP, SE and HM.

In Figure 2, we can observe that the AUC (95% CI) of model A is 0.744 (0.724-0.763). After incorporating optic disc parameters (VCDR, ovality index, cup-to-disc area ratio, rim area, disc area, and disc rotation) in the model, the AUC (95% CI) increased by 0.042 to 0.786 (0.768-0.805). The AUC comparison test revealed that the addition of optic disc parameters provided a significant improvement in predicting incident glaucoma (P <0.001).

**Figure 2.**
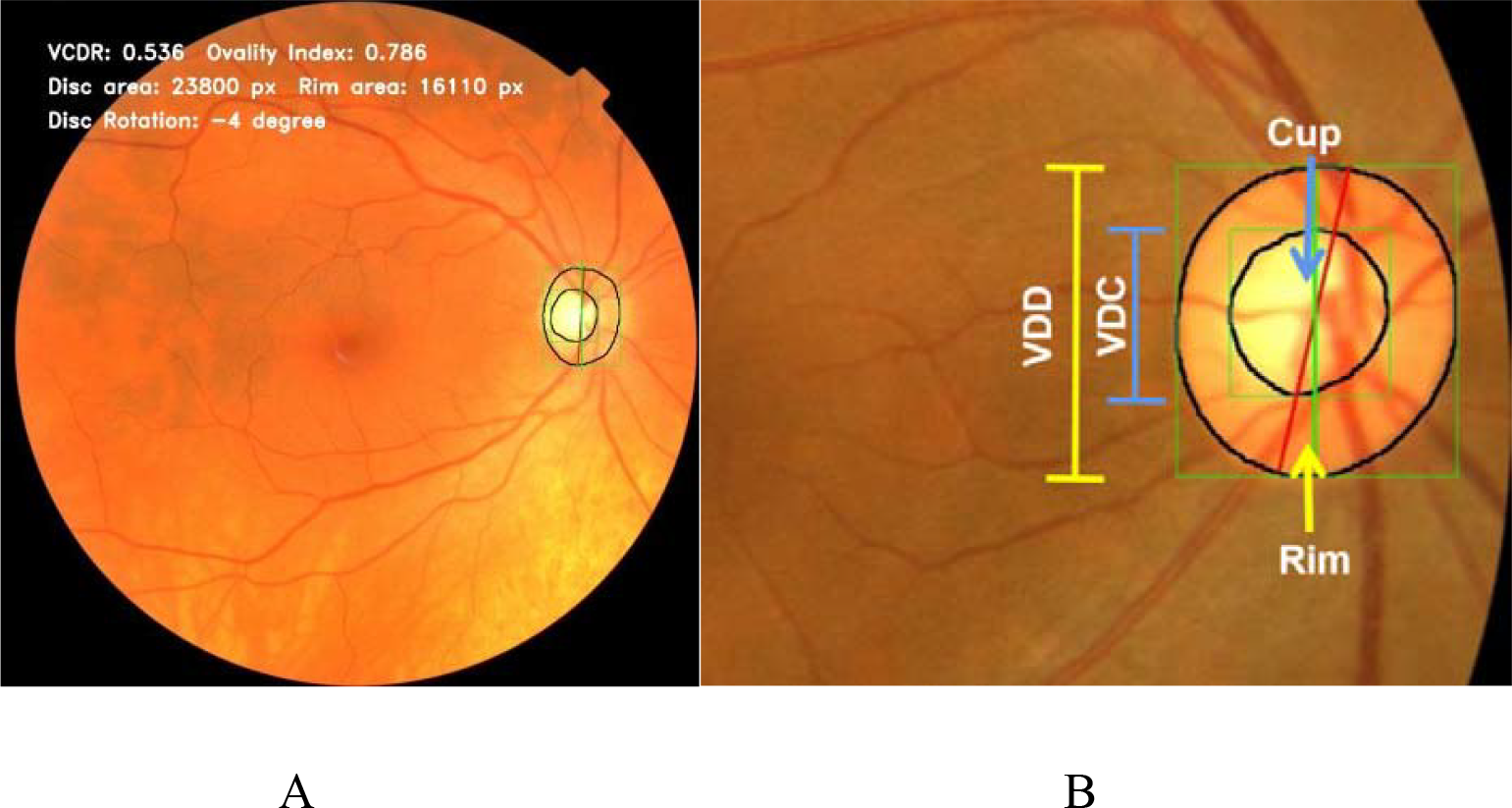
Schematic diagram of optic disc segmentation in CFPs. A: Example of segmentation results. B: Demonstration of disc-cup measurements. CFPs: Color Fundus Photos; VDD: Vertical Diameter of Disc; VDC: Vertical Diameter of Cup.

**Figure 3.**
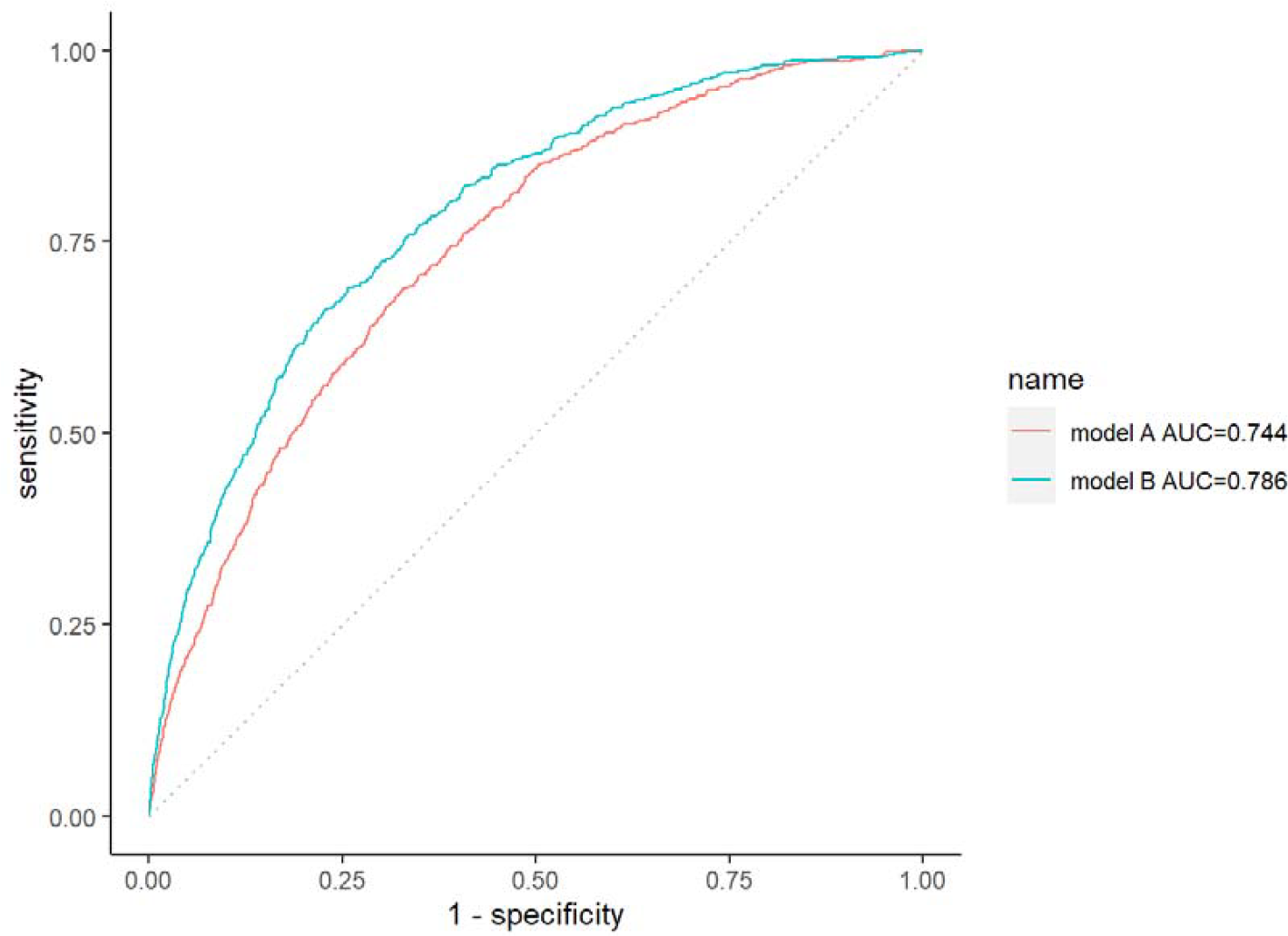
Receiver operating characteristic (ROC) curve of prediction models to the incidence of glaucoma. Model A was adjusted for age, sex, ethnicity, BMI, HDL, LDL, cholesterol, DBP, SBP, hypertension, diabetes, smoking, IOP, SE and HM; model B was additionally adjusted for VCDR, ovality index, cup to disc area ratio, rim area, disc area and disc rotation. BMI: Body Mass Index; SBP: Systolic Blood Pressure; DBP: Diastolic Blood Pressure; HDL: High-Density Lipoprotein; LDL: Low-Density Lipoprotein; IOP: Intraocular Pressure; SE: Spherical Equivalent; HM: High Myopia; VCDR: Vertical Cup-To-Disc Ratio.

## Discussion

In this study, our results indicate that the automatically segmented VCDR can be used as a reliable predictor for future risk of developing glaucoma. Our findings are consistent with previous studies^6,32,33^ that have shown a strong association between larger VCDR and glaucoma.

The changes in the optic disc in glaucoma are mainly reflected in the following aspects: progressive enlargement and deepening of the optic cup, significant thinning and narrowing of the superior and inferior rims, an increased cup-to-disc ratio in the vertical compared to horizontal direction, as well as superficial hemorrhage around the optic disc.^3,34,35^ According to research conducted at the Singapore National Eye Centre (SNEC), a CDR exceeding 0.65 indicates an increased risk of developing glaucoma.^7^ Gordon et al. demonstrated that a considerable cup-to-disc ratio in the vertical direction is a substantial predictor of primary open-angle glaucoma (POAG) development, which was further corroborated by Medeiros et al. in the population with ocular hypertension.^36,37^ Hence, despite being the most commonly used assessment for the diagnosis and monitoring of patients with suspected glaucoma, the clinical estimation of VCDR has limitations due to the physiological variability of this index in the population and the subjectivity of doctors’ judgments.^38,39^ Our findings revealed that the mean difference in VCDR between normal individuals and those at risk of glaucoma was only 0.07 (0.44 vs 0.51), indicating that it may be challenging for the human eye to accurately distinguish such a minor difference. Additionally, manual measurements of disc parameters, including VCDR, in previous studies have resulted in limited sample sizes, subjective variations, and challenges in standardization.

By incorporating the quantitative assistance of a DL model, clinicians will be better equipped to assess the risk of glaucoma from fundus photography that falls near the boundary value. A study conducted by Medeiros et al. demonstrated a hazard ratio (HR) of 1.32 for the development of glaucoma in an independent population of untreated subjects with ocular hypertension based on VCDR measurements.^37^ In comparison, our model showed a slightly higher HR of 2.05 for VCDR, which could be attributed to the predominance of healthy individuals in our study population.

The lack of statistical significance of other optic disc parameters, such as rim area, disc area, and cup-to-disc area ratio, in our study may be attributed to the fact that glaucoma primarily causes changes in the vertical diameter of the optic disc, and uniform enlargement of the optic cup may not be significantly associated with the risk of incident glaucoma, which is consistent with the underlying mechanism of glaucoma. Furthermore, a larger cup-to-disc area ratio is more likely to be associated with a physiological cup rather than glaucoma. Additionally, rim area and disc area exhibit high variability in the general population, and their sizes may not necessarily be directly linked to glaucoma development.^40^

In this study, we intentionally did not exclude fundus photographs with “Reject” quality, as we aimed to replicate the real-world scenario where image quality may vary. In our sensitivity analysis, we excluded these low-quality images and found that our results remained robust, indicating that the inclusion of images with varying quality did not significantly impact the findings.

This study has several notable strengths, including a large sample size, a long follow-up time, fully adjusted confounders, and the use of a DL model for automatic optic cup and disc segmentation. However, there are also several limitations that must be acknowledged. First, glaucoma was identified based on ICD10 coding records, self-reporting, and medical treatment records, potentially leading to the inclusion of undiagnosed cases. Second, the UK Biobank database primarily includes relatively healthy individuals aged 40-69 years, which may limit the generalizability of the findings to other populations. Third, our study mainly involved white individuals in Great Britain, and the results may not apply to other ethnicities or regions. Finally, the CPFs we used may have segmented optic discs inaccurately because of their inherent weaknesses.

## Data Availability

All data relevant to this manuscript will be available upon acceptance.

## Data availability statement

All data relevant to this manuscript will be available upon acceptance.

