## Supplemental Table 1 for "Association of optic disc parameters and glaucoma incidence based on automated segmentation, evidence from the UK Biobank"

**Table S1 Segmentation performance**

|  |  | **Accuracy** | **Sensitivity** | **Specificity** | **F1 score** |
| --- | --- | --- | --- | --- | --- |
| G1020 | Disc | 0.999 | 0.867 | 1.00 | 0.857 |
|  | Cup | 0.999 | 0.962 | 0.999 | 0.962 |
| GAMMA | Disc | 0.999 | 0.963 | 0.999 | 0.932 |
|  | Cup | 0.999 | 0.968 | 1.00 | 0.972 |
| REFUGE | Disc | 0.999 | 0.929 | 1.00 | 0.920 |
|  | Cup | 0.999 | 0.973 | 0.999 | 0.971 |
| RIGA | Disc | 0.999 | 0.830 | 0.999 | 0.808 |
|  | Cup | 0.999 | 0.976 | 0.999 | 0.962 |
| RIM-ONE | Disc | 0.998 | 0.937 | 0.999 | 0.905 |
|  | Cup | 0.998 | 0.971 | 0.999 | 0.976 |
| DRISHTI-GS | Disc | 0.999 | 0.938 | 0.999 | 0.947 |
|  | Cup | 0.999 | 0.977 | 0.999 | 0.978 |
