## Supplemental Table 2 for "Association of optic disc parameters and glaucoma incidence based on automated segmentation, evidence from the UK Biobank"

**Table S2. Association between optic disc parameters and incident glaucoma in sensitivity analyses.**

| **Optic disc parameters^γ^** | **Excluding 12,686 participants whose fundus photo quality was considered “Reject”**  **( n =31,684)** | | | | | |
| --- | --- | --- | --- | --- | --- | --- |
|  | **Model 1^*^** | | **Model 2^†^** | | **Model 3^‡^** | |
|  | **HR(95%CI)** | **P-value** | **HR(95%CI)** | **P-value** | **HR(95%CI)** | **P-value** |
| VCDR | 3.12(2.15-4.53) | <0.001 | 3.13(2.15-4.54) | <0.001 | 2.98(2.07-4.31) | <0.001 |
| Ovality Index | 1.08(0.92-1.28) | 0.335 | 1.08(0.92-1.28) | 0.338 | 1.08(0.92-1.27) | 0.370 |
| Disc Area (pixel) | 1.00(1.00-1.00) | 0.238 | 1.00(1.00-1.00) | 0.299 | 1.00(1.00-1.00) | 0.349 |
| Rim Area (pixel) | 1.00(1.00-1.00) | 0.114 | 1.00(1.00-1.00) | 0.141 | 1.00(1.00-1.00) | 0.239 |
| Cup To Disc Area Ratio | 0.58(0.33-1.02) | 0.060 | 0.61(0.35-1.07) | 0.084 | 0.66(0.37-1.17) | 0.153 |
| Disc Rotation | 1.01(1.00-1.01) | 0.202 | 1.00(1.00-1.01) | 0.224 | 1.00(1.00-1.01) | 0.278 |

BMI: Body Mass Index; SBP: Systolic Blood Pressure; DBP: Diastolic Blood Pressure; HDL: High Density Lipoprotein; LDL: Low Density Lipoprotein; IOP: Intraocular Pressure; SE: Spherical Equivalent; HM: High Myopia; VCDR:Vertical Cup-To-Disc Ratio; SD: Standard Deviation; CI: Confidence Interval; HR: Hazard Ratio.

^*^Model I adjusted for baseline age, sex, and ethnicity.

^†^Model II adjusted for baseline age, sex, ethnicity, BMI, HDL, LDL, Cholesterol, DBP, SBP, Hypertension, Diabetes and Smoking status.

^‡^Model III adjusted for baseline age, sex, ethnicity, BMI, HDL, LDL, Cholesterol, DBP, SBP, Hypertension, Diabetes, Smoking status, IOP, SE and HM.

^γ^Hazard ratios (HRs) and 95% confidence intervals (95%CIs) were calculated by Cox proportional hazards models.
